# Opioids Diminish the Placebo Antidepressant Response: A Post Hoc Analysis of a Randomized Controlled Ketamine Trial

**DOI:** 10.1101/2024.09.24.24314243

**Authors:** Theresa R. Lii, Josephine R. Flohr, Robin L. Okada, Lisa J. Cianfichi, Laura M. Hack, Alan F. Schatzberg, Boris D. Heifets

## Abstract

**Background:** The endogenous opioid system is thought to play a role in the placebo antidepressant response. A recent trial comparing the rapid antidepressant effects of ketamine versus placebo in surgical patients, some of whom were on chronic opioid therapy, revealed a substantial placebo effect. This finding provided an opportunity to test the hypothesis that opioid agonist exposure interacts with placebo antidepressant responses.

**Methods:** This *post hoc* analysis utilized data from a previously reported randomized, anesthesia-blinded, placebo-controlled trial of intravenous ketamine in depressed patients undergoing routine surgery. Mixed-effects models were used to determine whether baseline opioid use influenced antidepressant responses to the trial interventions, as measured by the Montgomery-Åsberg Depression Rating Scale (MADRS) over 1 to 14 days post-treatment.

**Results:** In the placebo arm, baseline opioid use was associated with a 10-point increase (95% CI: 0.81–19.4) in MADRS scores across all post-treatment time points, indicating worse depression in this subgroup. In an alternative model using percent change in MADRS scores, the difference between opioid users and non-users was 38.4% (95% CI: 8.59–68.2), with opioid users experiencing less improvement. For ketamine-treated participants, baseline opioid use did not significantly impact MADRS scores or the percent change in MADRS scores. Pain intensity was not a significant predictor of MADRS outcomes, and the correlation between post-treatment MADRS scores and pain intensity was negligible (R=0.12).

**Limitations:** This analysis was unregistered and conducted on a small sample; the findings need to be confirmed by prospective controlled studies.

**Conclusions:** Opioid use at baseline attenuated the placebo antidepressant response independently of pain in depressed patients who received the study treatment under general anesthesia for routine surgery. The antidepressant response was preserved in opioid users who received intravenous ketamine.

## 1. Introduction

The endogenous opioid system plays a critical role in regulating mood, reward, and emotional responses, making it a significant area of interest in depression research(Peciña et al., 2019). Alterations in endogenous opioid signaling may contribute to the pathophysiology of depressive disorders(Jelen et al., 2023; Peciña et al., 2019), highlighting a potential new target for therapeutic interventions. Moreover, the endogenous opioid system has been implicated in the modulation of placebo responses(Atlas et al., 2012; Benedetti et al., 2023; Colloca et al., 2013; Peciña et al., 2021; Wager and Atlas, 2015), particularly in the context of antidepressant trials, where placebo effects can be robust and clinically relevant(Jones et al., 2021).

Chronic opioid therapy, often used in the management of chronic pain and opioid use disorder, might induce changes in the endogenous opioid system that potentially impact mood(Ballantyne and Sullivan, 2017; Higginbotham et al., 2022). Chronic exposure to opioid agonists leads to adaptations at the receptor level, such as receptor downregulation or changes in signaling pathways(Corder et al., 2018), which could alter the natural capacity of the endogenous opioid system to regulate mood. The interaction between opioid use and the endogenous opioid system in the context of depression remains under-explored and represents a gap in our current understanding of this disease. Addressing this gap could significantly impact how we manage and study depression, particularly in patients on chronic opioid therapy.

We recently reported on a double-blind, placebo-controlled trial of ketamine administered during surgical anesthesia, which found a substantial placebo response comparable to the effect of ketamine observed in prior studies and in our trial(Lii et al., 2023). Here, we explored whether baseline opioid use status impacted the antidepressant effects of placebo and ketamine administration, allowing us to generate hypotheses about the relationship between the endogenous opioid system and the placebo antidepressant response.

## 2. Methods

### 2.1. Study Design and Participants

This was a non-registered, *post hoc* analysis of a single-center, double-blind, parallel-arm, randomized controlled trial which enrolled 40 adults with major depressive disorder (MDD) scheduled for routine surgery. Participants were randomized 1:1 to receive a single infusion of ketamine (0.5 mg/kg over 40 minutes) or a placebo (saline) during general anesthesia for surgery. For a complete description of the study design and eligibility criteria, please refer to the primary manuscript(Lii et al., 2023). The study protocol was pre-registered (NCT03861988) and approved by Stanford University’s institutional review board. All participants provided written informed consent.

### 2.2. Assessments

The pre-registered primary outcome of the original study was depression severity, measured by the Montgomery-Åsberg Depression Rating Scale (MADRS) across post-treatment days 1, 2, and 3. MADRS scores were also obtained on days 5, 7, and 14 for exploratory purposes. As a sensitivity measure, the Hospital Anxiety and Depression Scale (HADS) was also administered concurrently with the MADRS. For this *post hoc* analysis, participants were classified as opioid users at baseline if they reported using prescription opioid agonists during screening, confirmed with the state prescription drug monitoring database. Of note, patients using >90 morphine milligram equivalents (MME) per day, opioid antagonists, or partial agonists were excluded from the original study. Pain intensity was measured using the “pain on average” item in the Brief Pain Inventory, which is self-rated on a 0-10 numeric scale.

### 2.3. Statistical Analyses

#### 2.3.1. Model Selection

For each trial arm (ketamine vs. placebo), we used linear mixed-effects models with unstructured variance-covariance structures to analyze the impact of baseline opioid use status on MADRS scores from all post-treatment time points up to day 14. We employed a forward selection approach to build a model with additional variables to control for potential confounders (**Table 1**). The minimal model included subject ID as a random effect and group (opioid user vs. non-user), time, and group-by-time interaction as fixed effects. Additional variables were added iteratively based on their theoretical importance. We included baseline MADRS scores to account for the influence of initial depression severity. We included baseline and postoperative pain intensity as the bidirectional relationship between pain and mood is well-documented(Kroenke et al., 2011; Leuchter et al., 2010). We also included ethnicity since we observed a significant imbalance in Hispanic ethnicity between opioid users and non-users in our study (**Table 2**), and it has been suggested that race and ethnicity can influence the placebo response(Friesen and Blease, 2018). For analysis purposes, the one patient who withheld their ethnicity was classified as non-Hispanic.

**Table 1.** Forward Selection of Linear Mixed-Effects Models. Forward selection of linear mixed-effects models for post-treatment MADRS scores and percent change in MADRS scores in the placebo arm, highlighting the impact of additional predictors on model fit as indicated by AIC values.

| **Table 1. Forward Selection of Linear Mixed-Effects Models** |  |
| --- | --- |
| **For post-treatment MADRS scores (placebo arm)** |  |
| **Minimal model** | **AIC** |
| [Random effect]+[opioid use status]+[time]+[opioid use status*time] | 778.22 |
| **Minimal model with additional predictors** |  |
| [Min. model]+[baseline MADRS score] | 776.73 |
| [Min. model]+[baseline MADRS score]+[baseline pain intensity] | 698.72 |
| [Min. model]+[baseline MADRS score]+[baseline pain intensity]+[postop pain intensity] | 683.66 |
| [Min. model]+[baseline MADRS score]+[baseline pain intensity]+[postop pain intensity]+[ethnicity] | 678.38 |
| **For the % change in MADRS scores (placebo arm)** |  |
| **Minimal model** | **AIC** |
| [Random effect]+[opioid use status]+[time]+[opioid use status*time] | 1041.60 |
| **Minimal model with additional predictors** |  |
| [Min. model]+[baseline pain intensity] | 933.48 |
| [Min. model]+[baseline pain intensity]+[postop pain intensity] | 911.28 |
| [Min. model]+[baseline pain intensity]+[postop pain intensity]+[ethnicity] | 903.33 |

AIC, Akaike Information Criterion, a measure of fit of the model where smaller values represent better fit.

**Table 2.** Baseline Demographics and Clinical Characteristics. Baseline demographics and clinical characteristics of participants using and not using opioids at baseline, including comparisons of age, sex, race, ethnicity, marital status, body mass index, MDD characteristics, treatment history, and symptom severity scores.

| **Characteristic** | **Using**<br>**opioids at**<br>**baseline**<br>**(N=16)** | **Not using**<br>**opioids at**<br>**baseline**<br>**(N=24)** | **p value** |
| --- | --- | --- | --- |
| Age — yr | 55.9 ± 13.0 | 48.0 ± 16.7 | 0.103 |
| Female sex — no. (%) | 10 (63) | 18 (75) | 0.398 |
| White race — no. (%) | 10 (63) | 17 (71) | 0.582 |
| Hispanic ethnicity — no. (%) | 4 (25) | 0 (0) | 0.02† |
| Married — no. (%) | 7 (44) | 11 (46) | 0.897 |
| Body mass index — kg/m<sup>2</sup> | 30.3 ± 4.1 | 29.3 ± 5.1 | 0.515 |
| Age at first MDD onset — yr | 30.2 ± 13.9 | 24.2 ± 15.8 | 0.225 |
| Duration of current MDD episode — months | 53.8 ± 89.4 | 67.4 ± 93.5 | 0.647 |
| Median values | 21 | 24 |  |
| Recurrent MDD — no. (%) | 9 (56) | 18 (75) | 0.215 |
| Antidepressant augmentation trialed — no. (%) | 3 (19) | 7 (29) | 0.711† |
| Electroconvulsant therapy trialed — no. (%) | 0 (0) | 0 (0) | 1.000 |
| Maudsley Staging Method score | 7.4 ± 1.4 | 8.2 ± 2.3 | 0.168 |
| Maudsley Staging Method resistance category — no. (%) |  |  |  |
| Mild (<7) | 4 (25) | 5 (21) | >0.999† |
| Moderate (≥7 and <11) | 12 (75) | 15 (63) | 0.503† |
| Severe (≥11) | 0 (0) | 4 (17) | 0.136† |
| MADRS total score | 28.4 ± 5.5 | 29.6 ± 8.1 | 0.577 |
| HADS |  |  |  |
| Total score | 23.6 ± 6.1 | 25.2 ± 5.6 | 0.399 |
| Depression subscore | 10.8 ± 3.3 | 11.5 ± 3.6 | 0.477 |
| Anxiety subscore | 12.9 ± 3.8 | 13.7 ± 4.0 | 0.511 |
| Pain intensity rating | 6.1 ± 2.3 | 4.5 ± 2.4 | 0.059 |

†Fisher's exact test was used for cell counts <5. Otherwise, chi-squared test was used to compare proportions. Unpaired t test was used to compare means. Plus-minus values represent mean ± standard deviation. Participants were defined as using opioids if they self-reported taking prescription opioids during screening, corroborated by prescription drug monitoring database queries. Percentages may not total 100 due to rounding. Abbreviations: MDD, major depressive disorder; MADRS, Montgomery-Åsberg Depression Rating Scale; HADS, Hospital Anxiety and Depression Scale.

We evaluated the model fit using the Akaike Information Criteria (AIC); we compared the AIC of the current model with the AIC of the model with the newly added predictor. If adding a variable resulted in a lower AIC, indicating an improved model fit, the variable was retained in the model. The final model included subject ID as a random effect and group, time, group-by-time interaction, baseline MADRS score, baseline pain intensity, postoperative pain intensity, and ethnicity as fixed effects. Multicollinearity was assessed using the variance inflation factor (VIF), and all variables had VIF<5, indicating that multicollinearity was minimal. We also used the final model to analyze the percent change in MADRS scores relative to screening values; this model omitted baseline MADRS scores as a variable, since baseline scores are already accounted for in change scores. Due to the exploratory nature of this analysis, corrections for multiplicity were not performed.

#### 2.3.2. Sensitivity Analyses

The following sensitivity analyses were each conducted separately: (1) using the model to predict HADS scores instead of MADRS scores; (2) replacing baseline opioid use status in the model with the baseline self-reported daily dose of opioids. To evaluate the correlation between postoperative pain and depression severity, we conducted Pearson’s correlation analysis between postoperative pain intensity scores and MADRS scores for all post-treatment time points. All analyses were performed using RStudio software (version 2023.12.1 for MacOS). The lme4 package was used for mixed-effects modeling. Figures were made using Prism GraphPad (version 10.2.3 for MacOS).

## 3. Results

### 3.1. Baseline Characteristics

Baseline characteristics of opioid-users and non-users are compared in **Table 2**. Overall, clinical characteristics were similar between opioid users and non-users at baseline. However, there was a meaningful difference in self-reported Hispanic ethnicity in which 25% of opioid users identified as Hispanic, compared to 0% of non-users (p=0.02 by Fisher’s exact test). Hispanic ethnicity was therefore included as a fixed effect in our models. Prescription opioid use was reported by 45% and 35% (p=0.52) of participants in the ketamine and placebo arms, respectively, at screening. Self-reported daily opioid dose, in MME, at screening was similar between treatment arms (9.1 for ketamine; 10.5 for placebo; CI of the difference: −16.1 to 13.3).

### 3.2. Impact of Baseline Opioid Use on Antidepressant Response

**Figure 1.** illustrates MADRS scores (A,B) and percent change in MADRS scores (C,D) over time, grouped by treatment arm and baseline opioid use status; we observed that opioid users have a reduced antidepressant response to placebo—but not to ketamine. The mixed-effects model for placebo-only data revealed that baseline opioid use status and baseline MADRS scores significantly influenced the placebo antidepressant response (p=0.03 and p=0.02, respectively). Specifically, opioid use at baseline was associated with a 10-point increase (95% CI: 0.81-19.4) in MADRS scores across all post-treatment time points in the placebo arm. The significant effect of baseline opioid use status persisted (p=0.01) in the mixed-effects model predicting the percent change in MADRS scores; in this model, opioid use at baseline resulted in a 38.4% (95% CI: 8.59 to 68.2) reduction in the percent change in MADRS scores across all post-treatment time points. For participants who received ketamine, baseline opioid use did not significantly impact post-treatment MADRS scores or percent change in MADRS scores (p=0.50 and p=0.84, respectively). See **Supplemental Table 1** for model estimates.

**Figure 1.**
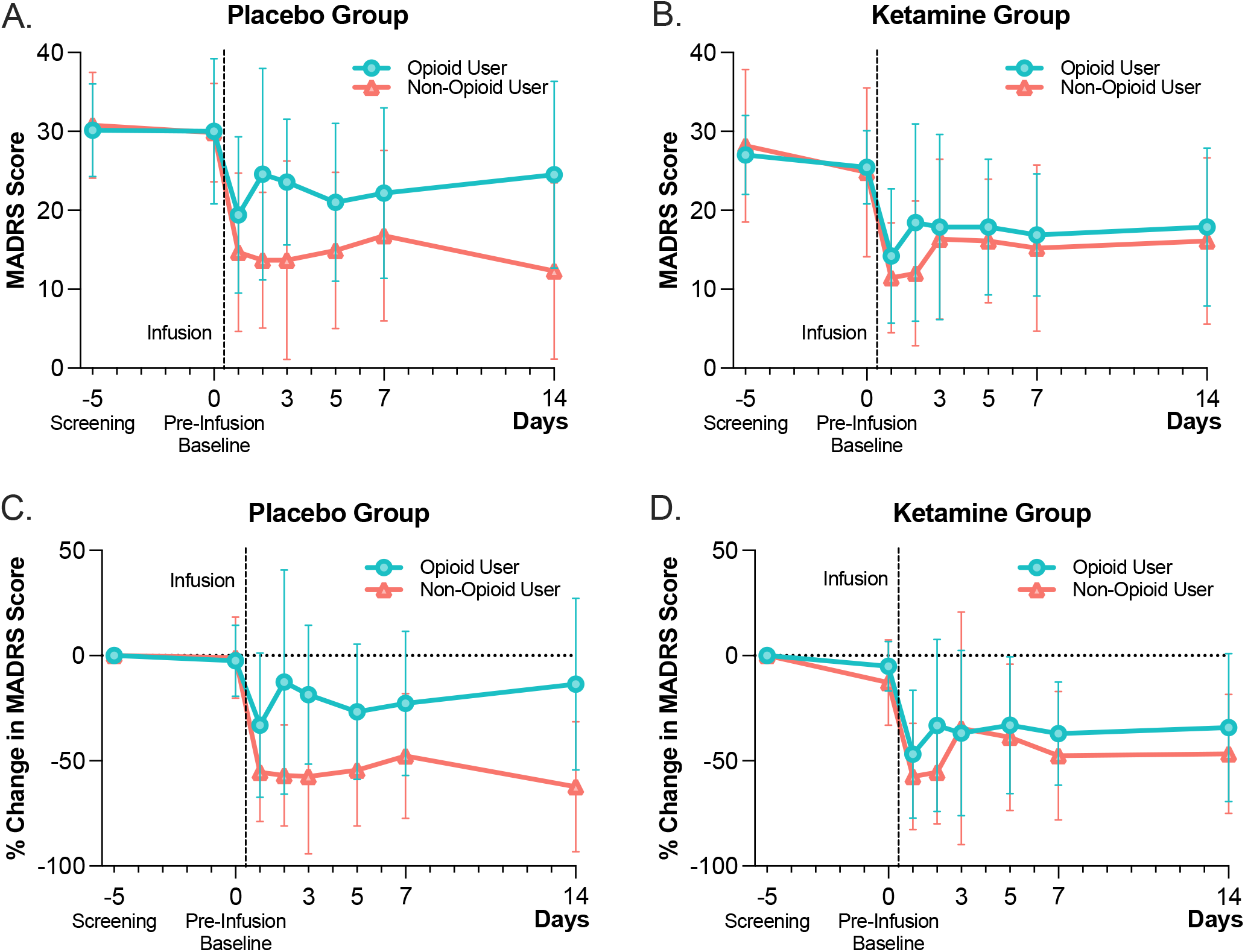
Baseline Opioids Reduced Antidepressant Response to Placebo, Not Ketamine. In a randomized, anesthesia-blinded, placebo-controlled trial of intravenous ketamine, baseline opioid use was associated with a smaller antidepressant response to placebo (A, C), whereas the antidepressant response to ketamine (B, D) was relatively preserved in opioid users. Mean and standard deviation scores are displayed. MADRS scores range from 0 to 60, with higher scores indicating greater depression. The percent (%) change in MADRS scores is relative to the MADRS score obtained during the screening visit. For simplicity of illustration, the screening visit is represented on day −5 for all participants since the screening visit took place on average 5 days before the infusion.

### 3.3. Relationship with Pain

Baseline and postoperative pain intensity were not significant predictors of post-treatment MADRS scores or percent change in MADRS scores in our mixed-effects models (**Supplemental Table 1**). The correlation between post-treatment MADRS scores and postoperative pain intensity was also negligible (Pearson’s R=0.12); although, as expected, patients reporting opioid use at baseline experienced more postoperative pain (visualized in **Figure 2**; mean[SD] values for pain intensity measured at all post-treatment time points: 6.0[2.2] vs. 4.5[1.9]; p<0.001).

**Figure 2.**
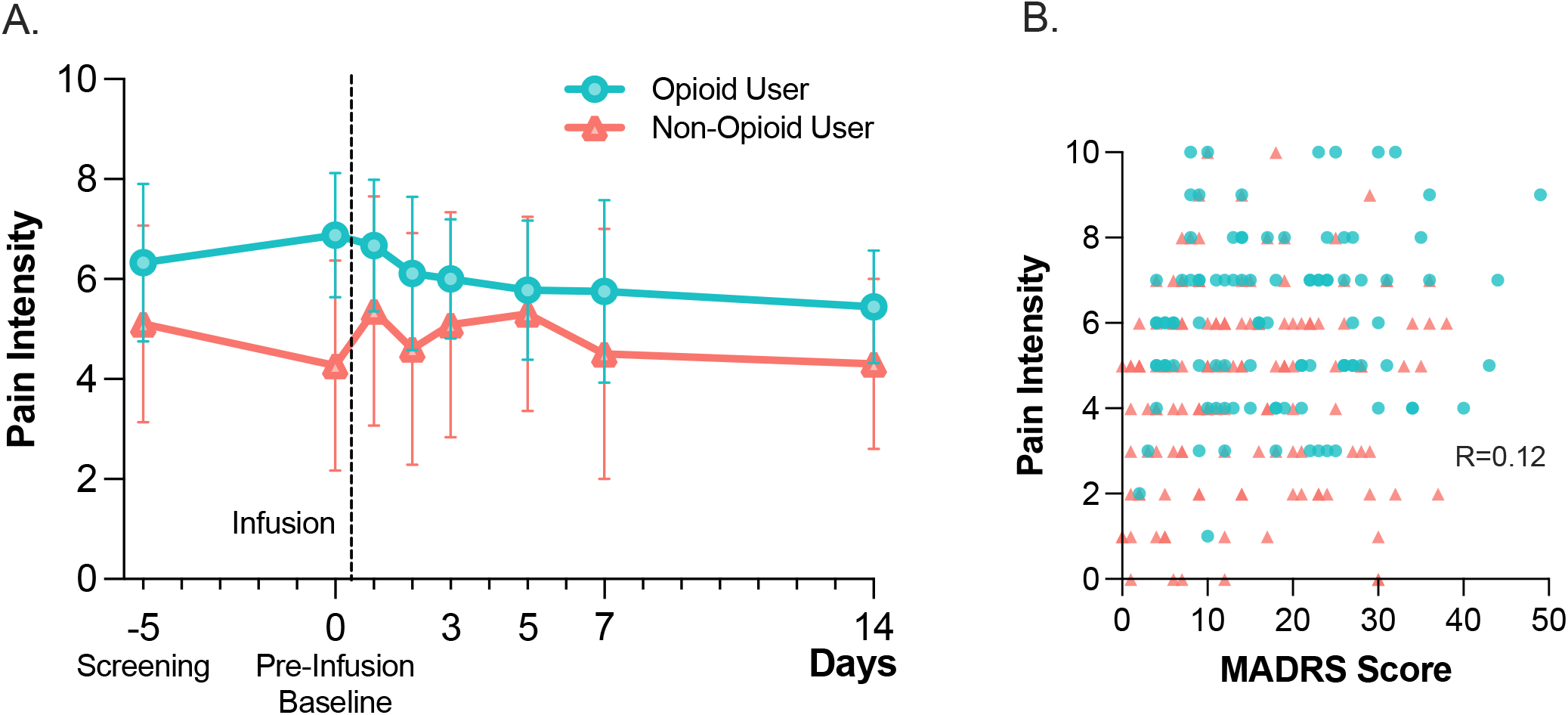
Relationships Between Baseline Opioid Use, Pain, and Depression Severity. (A) As expected, opioid users at baseline (blue circles) experienced more pain across the entire study compared to non-users (red triangles). Mean and standard deviations are shown. Pain intensity scores are self-rated on a 0-10 numeric rating scale. (B) There is negligible correlation between postoperative pain intensity and MADRS scores (Pearson’s R=0.12 for all post-treatment data points, including opioid users [blue circles] and non-users [red triangles]).

### 3.4. Sensitivity Analyses

We tested whether our results were sensitive to an alternative measure of mood: the HADS, which unlike the MADRS, relies on self-report and de-emphasizes somatic symptoms. Using our final model to predict total HADS scores, the fixed effects of time and the interaction between time and baseline opioid use status were significant in the placebo group (p<0.001 and p=0.01, respectively). The effect of baseline opioid use status on HADS scores increased significantly over time: the difference grew by 0.41 points per day (95% CI: 0.08 to 0.74; visualized in **Supplemental Figure 1**). Independently of time, however, the impact of baseline opioid use status was not significant, due to a smaller difference in HADS scores observed during the early postoperative period. Similar findings were obtained using our final model to predict the percent change in HADS scores (**Supplemental Table 1**). To evaluate whether a dose-response relationship exists for opioid use, we replaced baseline opioid use status—a binary variable—in our final model with the baseline self-reported opioid dose in MME/day; this was not a significant predictor of post-treatment MADRS scores (p=0.30) or percent change in MADRS scores (p=0.19).

## 4. Discussion

### 4.1. Summary

In this *post hoc* analysis of a randomized, anesthesia-blinded, placebo-controlled trial of ketamine in adults with MDD, we observed that baseline opioid use status reduced the placebo antidepressant response independently of pain, and that the antidepressant effect of ketamine was unaffected by baseline opioid use. Our analysis controlled for baseline depression severity, baseline pain intensity, postoperative pain intensity, and ethnicity. In sensitivity analyses, an alternative measure of mood (HADS) showed that the attenuating effect of opioid use on antidepressant response significantly increased over time in the placebo arm. No dose-response relationship was found between baseline self-reported opioid dose and the reduction in the placebo antidepressant effect.

### 4.2. Limitations

First, the reliability of our findings are limited by the *post hoc* nature of this analysis, as well as the small sample size (9 opioid users in the ketamine arm, 7 opioid users in the placebo arm) which increases the risk of a type I error. These findings should, therefore, be interpreted cautiously and confirmed by prospective randomized placebo-controlled studies. Second, we did not assess patients’ adherence to their self-reported opioid dose nor the proximity of their last opioid dose to the trial intervention. The lack of adherence data and timing data may have obscured the dose-response relationship between baseline opioid dose and the placebo antidepressant response. Third, despite controlling for baseline depression severity, pain, and ethnicity, there may be unmeasured confounders related to baseline opioid use which might reduce the placebo antidepressant response (e.g. polymorphisms in the µ-opioid receptor gene associated with chronic pain(Peciña et al., 2019)).

### 4.3. Follow-On Hypotheses & Future Directions

Based on our findings, we hypothesize that exogenous opioids might disrupt the endogenous opioid system’s ability to mount a placebo antidepressant response. Despite disproportionately high rates of prescription opioid use among depressed individuals(Davis et al., 2017), the impact of chronic opioid exposure in antidepressant trials has been surprisingly under-explored. Previous research by Stahl *et al*. indicated that opioid use diminishes analgesia from venlafaxine without affecting its antidepressant effects; however, their study did not include a placebo condition to assess the influence of opioid use on placebo responses(Stahl et al., 2020).

We also hypothesize that the unchanged response to ketamine in opioid users is explained by ketamine’s ability to reverse opioid tolerance(Kissin et al., 2000; Laskowski et al., 2011), enabling a full antidepressant response to endogenous—or exogenous—opioids, despite recent opioid use. This hypothesis aligns with findings that naltrexone, an opioid antagonist, can block ketamine’s antidepressant effects(Williams et al., 2018), highlighting the key role of opioid signaling in ketamine’s mechanism of action.

Future research should investigate the extent to which chronic opioid agonist and antagonist therapy affects placebo responses in antidepressant trials. We also advocate for the use of opioid antagonists, such as naltrexone, as tools to probe the placebo response in mechanistic trials. If the mechanisms underlying rapid-acting antidepressants like ketamine and psilocybin overlap with those of placebo, enhancing sensitivity to placebo could be a viable strategy for developing new therapeutics for various neuropsychiatric conditions.

## Data Availability

De-identified participant data, accompanying data dictionaries, the study protocol, and the statistical analysis plan are accessible at https://osf.io/zdkr8/. All participants provided consent for their de-identified data to be shared with external entities for scientific research purposes.

## Acknowledgements

We would like to thank Ashleigh Smith, Cynthia Nyongesa, Kayla Pfaff, and Rasmus Thordstein for their valuable assistance in screening, recruitment, and participant follow-up.

