## Supplemental Figure 1 for "Opioids Diminish the Placebo Antidepressant Response: A Post Hoc Analysis of a Randomized Controlled Ketamine Trial"

**Supplmental Figure 1.**

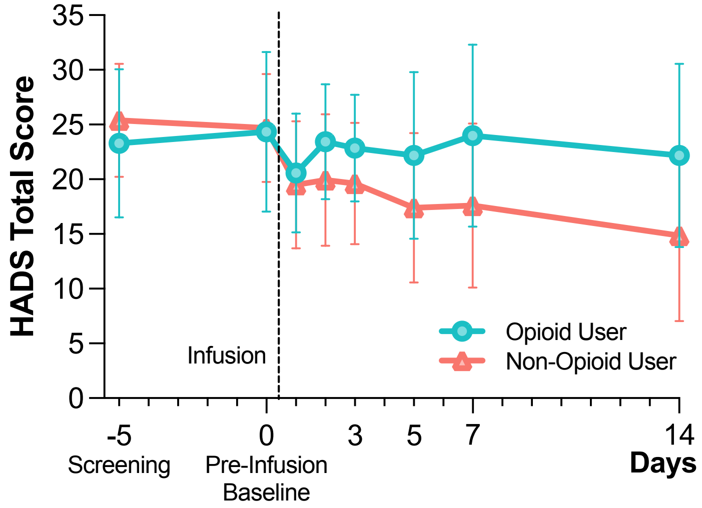


**Caption for Supplmental Figure 1.**

**Impact of Baseline Opioid Use on HADS Scores in the Placebo Group**

Baseline opioid use was associated with a smaller antidepressant response to placebo, as measured by the HADS (Hospital Anxiety and Depression Scale), a secondary self-report measure of mood that de-emphasizes somatic symptoms and is specifically validated on hospitalized patients. Mean and standard deviation scores are displayed. HADS total scores range from 0-42, with higher scores indicating greater anxiety and depression symptoms.
