## Supplemental Table 1 for "Opioids Diminish the Placebo Antidepressant Response: A Post Hoc Analysis of a Randomized Controlled Ketamine Trial"

| Supplemental Table 1. Model Estimates | | | | |
|---|---|---|---|---|
| **Model for post-treatment MADRS scores (placebo arm)** | | | | |
| | **Estimate (SE)** | **95% CI** | **t-value** | **p-value** |
| Intercept | -2.86 (10.1) | -22.7 to 17.0 | -0.28 | 0.777 |
| Group (baseline opioid use status) | 10.1 (4.73) | 0.81 to 19.4 | 2.13 | 0.033* |
| Time | -0.07 (0.17) | -0.40 to 0.27 | -0.39 | 0.699 |
| Group×Time | 0.24 (0.29) | -0.32 to 0.79 | 0.83 | 0.409 |
| Baseline MADRS score | 0.70 (0.31) | 0.09 to 1.31 | 2.26 | 0.024* |
| Baseline pain intensity | -1.34 (0.74) | -2.78 to 0.11 | -1.81 | 0.070 |
| Postoperative pain intensity | 0.50 (0.42) | -0.33 to 1.33 | 1.18 | 0.239 |
| Ethnicity | -9.21 (9.22) | -27.3 to 8.87 | -1.00 | 0.318 |
| **Model for % change in MADRS scores (placebo arm)** | | | | |
| | **Estimate (SE)** | **95% CI** | **t-value** | **p-value** |
| Intercept | -43.8 (12.7) | -68.7 to -19.0 | -3.46 | 0.001* |
| Group (baseline opioid use status) | 38.4 (15.2) | 8.59 to 68.2 | 2.53 | 0.012* |
| Time | -0.15 (0.56) | -1.25 to 0.95 | -0.27 | 0.789 |
| Group×Time | 0.54 (0.95) | -1.33 to 2.41 | 0.57 | 0.570 |
| Baseline pain intensity | -3.81 (2.40) | -8.51 to 0.89 | -1.59 | 0.112 |
| Postoperative pain intensity | 1.75 (1.42) | -1.02 to 4.52 | 1.24 | 0.216 |
| Ethnicity | -34.4 (30.0) | -93.1 to 24.3 | -1.15 | 0.251 |
| **Model for post-treatment MADRS scores (ketamine arm)** | | | | |
| | **Estimate (SE)** | **95% CI** | **t-value** | **p-value** |
| Intercept | -2.30 (7.15) | -16.3 to 11.7 | -0.32 | 0.748 |
| Group (baseline opioid use status) | 2.63 (3.87) | -4.96 to 10.2 | 0.68 | 0.497 |
| Time | 0.31 (0.20) | -0.09 to 0.70 | 1.52 | 0.127 |
| Group×Time | -0.11 (0.28) | -0.67 to 0.44 | -0.40 | 0.686 |
| Baseline MADRS score | 0.50 (0.23) | 0.05 to 0.95 | 2.19 | 0.029* |
| Baseline pain intensity | -0.87 (1.22) | -3.26 to 1.52 | -0.72 | 0.474 |
| Postoperative pain intensity | 1.11 (0.63) | -0.13 to 2.34 | 1.75 | 0.079 |
| Ethnicity | 4.99 (6.14) | -7.03 to 17.0 | 0.81 | 0.416 |
| **Model for % change in MADRS scores (ketamine arm)** | | | | |
| | **Estimate (SE)** | **95% CI** | **t-value** | **p-value** |
| Intercept | -57.9 (21.5) | -100 to -15.7 | -2.69 | 0.007* |
| Group (baseline opioid use status) | 2.92 (14.5) | -25.5 to 31.3 | 0.20 | 0.840 |
| Time | 0.47 (0.84) | -1.18 to 2.11 | 0.55 | 0.579 |
| Group×Time | 0.29 (1.18) | -2.02 to 2.60 | 0.25 | 0.806 |
| Baseline pain intensity | -2.66 (4.25) | -11.0 to 5.68 | -0.62 | 0.532 |
| Postoperative pain intensity | 4.43 (2.58) | -0.63 to 9.49 | 1.72 | 0.086 |
| Ethnicity | 19.2 (22.5) | -24.9 to 63.4 | 0.85 | 0.393 |
| **Model for post-treatment HADS scores (placebo arm)** | | | | |
| | **Estimate (SE)** | **95% CI** | **t-value** | **p-value** |
| Intercept | 14.3 (6.93) | 0.71 to 27.9 | 2.06 | 0.039* |
| Group (baseline opioid use status) | 2.41 (3.60) | -4.64 to 9.46 | 0.67 | 0.502 |
| Time | -0.36 (0.10) | -0.55 to -0.16 | -3.61 | <0.001* |
| Group×Time | 0.41 (0.17) | 0.08 to 0.74 | 2.45 | 0.014* |

| | | | | |
|---|---|---|---|---|
| Baseline HADS | 0.24 (0.27) | -0.29 to 0.76 | 0.88 | 0.379 |
| Baseline pain intensity | -0.36 (0.58) | -1.50 to 0.78 | -0.62 | 0.533 |
| Postoperative pain intensity | 0.47 (0.25) | -0.02 to 0.96 | 1.90 | 0.058 |
| Ethnicity | -3.23 (7.06) | -17.1 to 10.6 | -0.46 | 0.648 |
| **Model for % change in HADS scores (placebo arm)** | | | | |
| | **Estimate (SE)** | **95% CI** | **t-value** | **p-value** |
| Intercept | 36.9 (25.2) | -12.6 to 86.3 | 1.46 | 0.144 |
| Group (baseline opioid use status) | 16.1 (13.2) | -9.80 to 42.1 | 1.22 | 0.223 |
| Time | -1.50 (0.45) | -2.38 to -0.63 | -3.36 | 0.001* |
| Group×Time | 1.52 (0.76) | 0.04 to 3.01 | 2.01 | 0.045* |
| Baseline pain intensity | -1.11 (2.13) | -5.28 to 3.06 | -0.52 | 0.601 |
| Postoperative pain intensity | 2.15 (1.12) | -0.05 to 4.34 | 1.92 | 0.055 |
| Ethnicity | -21.9 (25.7) | -72.1 to 28.4 | -0.85 | 0.394 |

*Statistical significance defined by p<0.05 without corrections. SE, standard error. CI, confidence interval. MADRS, Montgomery-Asberg Depression Rating Scale. HADS, Hospital Anxiety and Depression Scale. Pain intensity was rated on a 0-10 numeric scale. Baseline values were obtained during screening. Post-treatment values were obtained on postoperative days 1, 2, 3, 5, 7 and 14.

**Caption for Supplemental Table 1. Model Estimates**

Model estimates for post-treatment MADRS and HADS scores and percent change in scores for placebo and ketamine arms, detailing the effects of baseline opioid use status, time, and additional predictors on the outcomes.
